# Documenting HIV service delivery and adoption of evidence-based practices in HIV care programs: A protocol for a cross-sectional survey of HIV clinics participating in the International epidemiology Databases to Evaluate AIDS (IeDEA)

**DOI:** 10.64898/2026.09.16.26363245

**Authors:** Ellen Brazier, Fernanda Maruri, Stephany N. Duda, Kara Wools-Kaloustian, Aimee Freeman, Beverly Musick, Sophia Arabadjis, Marie Ballif, Nathalie Verónica Fernández Villalobos, Mark H. Kuniholm, Kathryn E. Lancaster, Sita Lujintanon, Angela M. Parcesepe, Rachael A. Pellegrino, Ana Fernanda Ramos Menchelli, Jeremy Ross, Jonathan Ross, Aggrey Semeere, Denis Nash

## Abstract

**Introduction:** HIV care and treatment programs around the world continue to evolve in response to changes in clinical guidelines, evidence-based practices, domestic and donor funding for the HIV response, new technological developments, and changing weather patterns. Since 2009, the International epidemiology Databases to Evaluate AIDS (IeDEA) research consortium has conducted periodic surveys to document the characteristics of participating clinics and the availability of HIV-related services across diverse settings—information that provides important context for understanding variations and trends in HIV care outcomes. This paper describes the protocol for IeDEA’s 2026 site assessment survey, launched in August 2026.

**Methods and analysis:** This cross-sectional descriptive study aims to document HIV-related services and practices among clinics in 42 countries. The survey explores the characteristics and services provided at HIV care and treatment sites participating in IeDEA in 2026, along with real-world adoption and implementation of evidence-based service delivery strategies for people living with or at risk of HIV. Designed for self-administration, the survey questionnaire was developed through two complementary processes: a review of questions and data collected through IeDEA’s prior consortium-wide surveys and an open call to IeDEA investigators for new survey content to advance a timely implementation science agenda relevant to the global HIV community. Survey materials and links to an online REDCap questionnaire will be distributed to 235 eligible clinics across IeDEA in September 2026, with a deadline of November 30, 2026, for completion. Planned analyses will examine the following topics: HIV care amidst funding constraints; the roll-out of differentiated service delivery models; pre-exposure HIV prophylaxis; long-acting injectable antiretroviral therapy (ART); prevention, screening and/or treatment services for viral hepatitis, mental health and substance use disorders, cardiovascular disease, and cancers among PLHIV; the integration of HIV care with other health services; and clinic vulnerability to extreme weather events.

**Ethics and dissemination:** A “non-human subjects” research designation was determined by the Vanderbilt University Medical Center Human Research Protections Program. Survey findings will be shared with local partners and stakeholders across IeDEA, disseminated at regional and global conferences, and published in peer-reviewed journals.

**Strengths and limitations of this study:**

- Strengths of this study include gathering data for a global cohort of 235 HIV clinics across 42 low-, middle- and high-income countries, reflecting real-world adoption and implementation of treatment guidelines and evidence-based practices for HIV prevention, care and management across diverse settings during a period of substantial changes in funding for the HIV response. Further strengths include a rigorous process for survey development that combined an in-depth review of data and findings from prior International epidemiology Databases to Evaluate AIDS (IeDEA) surveys with new investigator-driven content focused on implementation science questions related to the HIV response and key gaps in the literature.
- Study limitations include potential recall and social desirability biases inherent in self-reported data, as well as selection biases that may determine clinic participation. Additionally, while clinics participating in the global IeDEA research consortium are reflective of real-world HIV care delivery, they may not be nationally or regionally representative.

## I. Introduction

The past 25 years have witnessed extraordinary progress in addressing the global HIV epidemic through expanded access to life-saving treatment and prevention services, resulting in decreases in the number of new HIV infections, reductions in HIV-related mortality and rebounds in life expectancy in world regions most affected by HIV.^1^ These successes reflect the global scale up of evidence-based practices, along with improvements in HIV prevention and treatment and sustained efforts to improve outcomes along the HIV care continuum.

Alongside clinical trials, real-world service delivery data from observational cohorts, such as the International epidemiology Databases to Evaluate AIDS (IeDEA)^2^ research consortium have provided critical insights to guide the implementation of HIV treatment strategies and to monitor progress toward global treatment targets.^3 4^ Established in 2006 with funding from the U.S. National Institutes of Health to collect and harmonize observational data from clinical centers and research groups serving people living with and at risk for HIV, IeDEA has comprised close to 400 HIV clinics and programs across 44 countries in seven world regions.^2 5^ Reflecting HIV care outcomes in real-world service delivery settings operating under national treatment guidelines, IeDEA data have been used to address high priority and evolving research questions in HIV/AIDS treatment and care, ranging from global trends in the immunological status of people initiating antiretroviral therapy (ART) and late presentation to care among older individuals^6-10^ to the prevalence of infectious and non-infectious comorbid conditions among people living with HIV (PLHIV)^11-17^ and lingering sex and age disparities in the uptake of recommended first-line treatment regimens.^18-20^ IeDEA data have also provided important insights about care and treatment outcomes among groups generally not reflected in trial data, such as children, adolescents and pregnant women living with HIV,^21-23^ and informed mathematical modelling estimates of the HIV pandemic.^24-26^

Since 2009, IeDEA has conducted periodic surveys among sites participating in the consortium to document clinic characteristics, resources, and service provision to PLHIV. These surveys have helped describe the comprehensiveness of HIV care across the consortium,^27-29^ and real-world adoption and implementation of treatment guidelines and evidence-based practices, such as the scale-up up of the World Health Organization’s recommendations for universal HIV treatment,^30^ differentiated HIV service delivery,^31^ tuberculosis preventive therapy,^32^ management of HIV and tuberculosis co-infection,^33^ and country-level transition to dolutegravir-based ART regimens.^34^ IeDEA’s site surveys have provided snapshots of the availability of HIV-related services, including pre-exposure prophylaxis,^35^ mental health and substance use screening and treatment services for PLHIV,^36-39^ the integration of maternal health and HIV care,^40^ viral hepatitis prevention, screening and treatment,^41^ service delivery during the COVID-19 pandemic,^42 43^ cancer screening, prevention and care.^44 45^ An IeDEA site survey also provided early evidence of disruptions in HIV care after the freezing of U.S. foreign assistance funding in early 2025.^46^

Amidst increasing funding constraints for global health,^47 48^ restrictions on U.S. foreign assistance,^49-53^ reduced data on the HIV response^54^, and the variable burden of extreme weather hazards across world regions,^55^ observational cohorts, such as IeDEA, can provide a valuable window into the status of HIV-related service delivery across various levels of diverse national health systems and the implementation of evidence-based practices to optimize HIV care outcomes.^56^ Documenting the characteristics of clinics participating in IeDEA and the scope of services provided for PLHIV illuminates current clinic capacity and programmatic strategies across low- and high-resource settings, while providing important contextual information for interpreting IeDEA’s patient-level data and temporal trends in HIV care outcomes of interest to the global HIV community.

## II. Methods and Analysis

### Study setting

Since being established in 2006, IeDEA has harmonized patient data from HIV care and treatment programs in low-, middle- and high-income countries across seven geographic regions: the Asia-Pacific, Central Africa, East Africa, Southern Africa, West Africa, the Caribbean and Central/South America, and North America.^2^ Regional data centers receive regular data submissions from participating clinics in their respective regions, harmonize the data in accordance with IeDEA’s common data exchange standard^57^ and make it available for investigators with approved research proposals. Clinics participating in IeDEA include a mix of health care facilities providing HIV care and treatment, ranging from primary health centers in rural settings to tertiary referral and university teaching hospitals that provide specialist care.^5 58 59^ Clinics participating in IeDEA’s regional cohorts have changed over time, with some periodically exiting the research consortium and new ones joining.

### Study design and objectives

The study is the sixth cross-sectional self-administered survey of eligible IeDEA clinics since 2009. The overall objective of IeDEA’s site assessment survey is to document service availability and practices related to HIV prevention and care at clinics participating in IeDEA in 2026. This information is critical for characterizing trends in routine HIV care and specialized services for PLHIV across IeDEA in support of a timely scientific agenda of interest to the global HIV community. Specific objectives are to: (1) systematically document characteristics and services provided at HIV care and treatment sites participating in the consortium and describe temporal trends in HIV-related service delivery; and (2) collect data on specialized topics to address important scientific gaps in HIV research.

### Questionnaire development

The survey questionnaire was developed through two complementary processes, undertaken concurrently. First, questions fielded through IeDEA’s recent consortium-wide surveys in 2017, 2020 and 2023 were reviewed to identify questions that were no longer relevant given current HIV treatment guidelines. Additionally, data from these prior surveys were reviewed to identify questions with low response variability or potentially prone to social desirability biases or misinterpretation. Proposed cuts and changes to this legacy survey content were discussed during monthly calls of IeDEA’s Site Assessment Working Group (SAWG) to ensure broad agreement about these revisions.

Secondly, in accordance with the approach used to develop IeDEA’s 2020 and 2023 survey questionnaires,^60^ IeDEA’s SAWG issued a call for proposals, inviting IeDEA scientific working groups^61^ and affiliated investigators to prepare research proposals in support of new survey content. Investigators proposing new survey content were asked to articulate a formal research question and study objectives, along with specific survey questions and an analysis plan. The SAWG encouraged investigators to propose implementation science-focused research questions^62^ that could be explored via a self-administered survey. The SAWG circulated guidance to all IeDEA working groups and principal investigators leading IeDEA’s regional cohorts in early March 2026, with research proposals submitted through April 2026.

A total of 12 investigator-driven research proposals were submitted to IeDEA’s SAWG. Six members of the SAWG (EB, FM, DN, KW-K, SD, AF) reviewed and provided feedback to investigators on research aims and proposed survey questions. Eleven proposals moved forward and the SAWG worked with corresponding investigators to refine questions to minimize respondent burden, enhance the quality and interpretability of survey data, and standardize the formulation and wording of survey questions. Where relevant, new survey content was reviewed against and aligned with other reference survey instruments (e.g., World Health Organization’s Harmonized Health Facility Assessment [HHFA]^63^ and Service Availability and Readiness Assessment [SARA]),^64^ along with facility assessment tools related to non-communicable diseases^65^ and climate resilience.^66^

Following the approach used in prior IeDEA-wide general site assessment surveys, legacy and new survey content were compiled into a single questionnaire, with both a printable version and an online questionnaire in REDCap,^67^ hosted at Vanderbilt University Medical Center (VUMC). A printable questionnaire ensures that the survey can be completed in settings with intermittent internet access.

Additionally, the printed questionnaire can serve as a worksheet and/or be discussed with colleagues within a clinic, as needed, prior to completing the online survey.

The survey questionnaire includes 16 survey domains whose contents will be used in analyses focused on 11 research topics (Figure 1).

**Figure 1.**
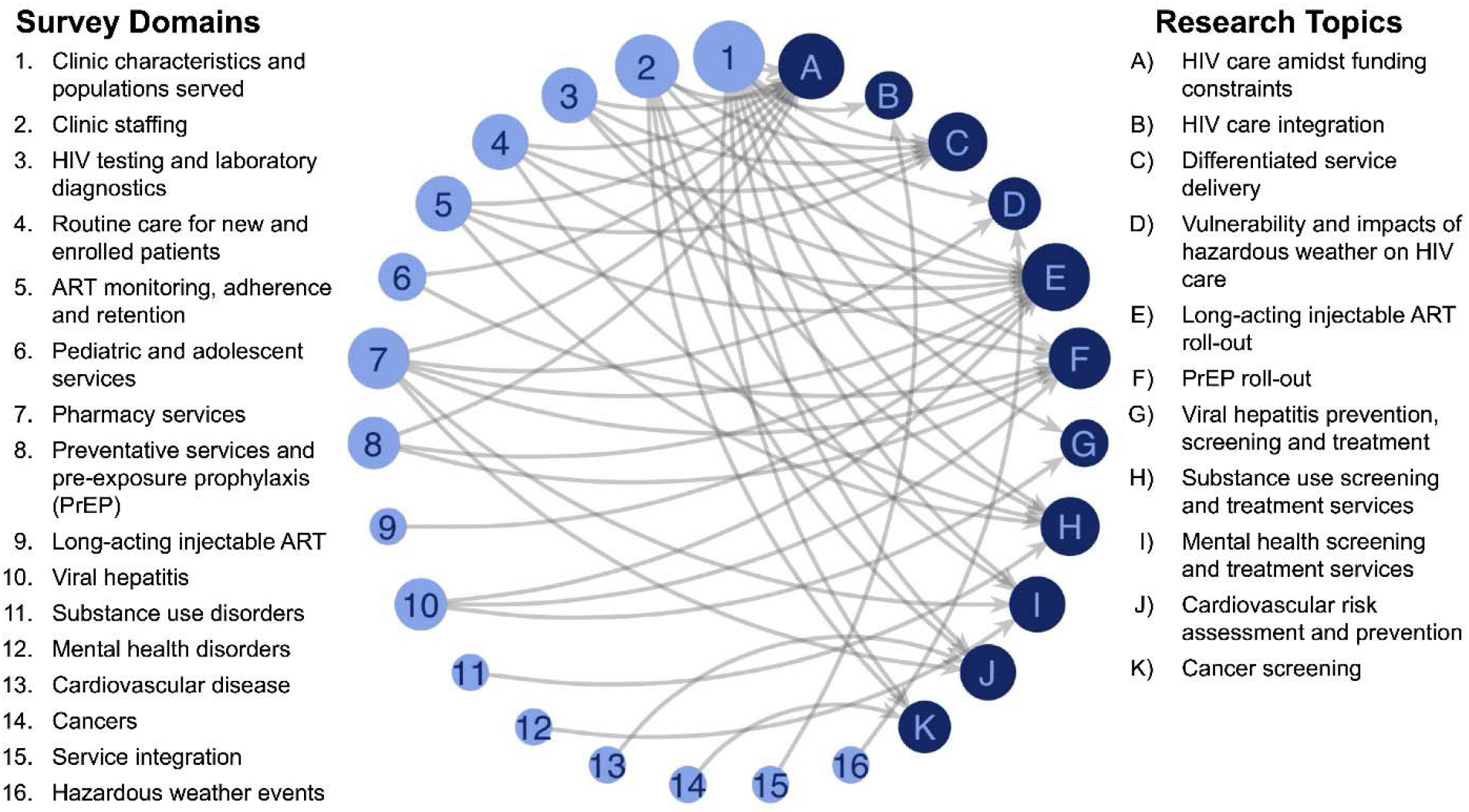
Survey domains and research topics to be explored through IeDEA’s 2026 general site assessment survey. ***Legend***. *A visual summary of survey domains (light blue) and the research topics (dark blue) for IeDEA’s 2026 site assessment survey. Lines indicate connections between survey domains and research topics. Circle size reflects the number of research topics linked to each survey domain and the number of survey domains linked to each research topic*.

### Survey piloting

IeDEA regions were asked to identify up to two clinics that could participate in piloting the survey. Pilot sites were asked to complete the survey within a two-week period in July 2026 and to provide feedback on the amount of time required to complete the survey and on any unclear questions or instructions. Ten clinics across eight countries in six IeDEA regions were invited to participate in the pilot; seven clinics in six countries (Figure 2) completed the pilot survey by the deadline, including one primary health center and six tertiary referral hospitals. Their responses were reviewed for logical consistency and potential social desirability biases, and the formulation of questions was revised to address identified concerns. The finalized questionnaire (Supplementary material 1) was then translated into French, Portuguese and Spanish and deployed in REDCap in all four languages.

**Figure 2.**
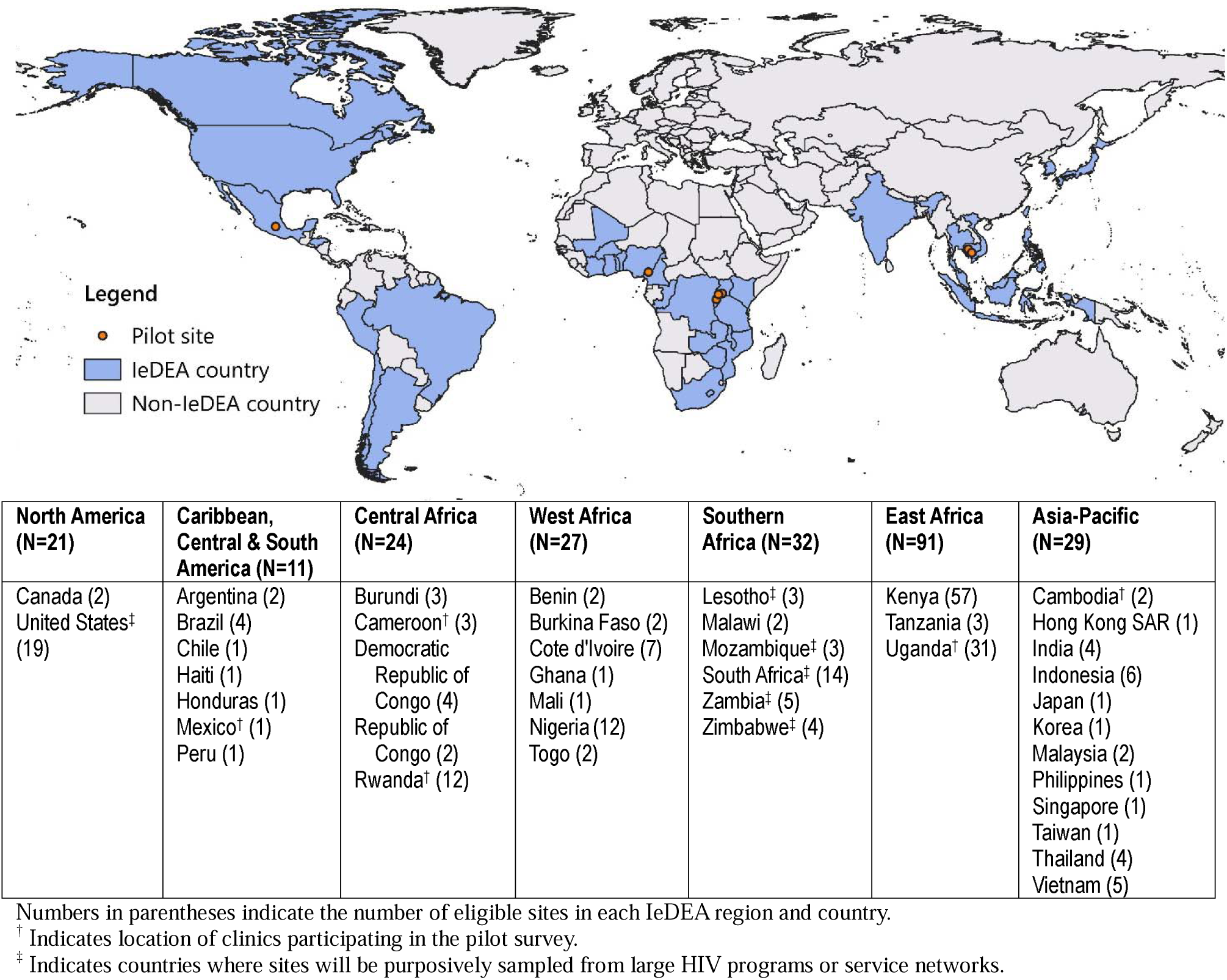
Map of countries with clinics eligible for IeDEA’s 2026 site assessment survey and sites piloting the survey (n=7)

### Study sample

Because clinics periodically enter and exit the consortium, each IeDEA region was requested to review and update their listing of sites in IeDEA’s central database of clinics ever participating in the consortium. All active clinical sites providing data from routine patient visits directly to IeDEA’s country or regional data centers in 2026 were eligible for the survey. In IeDEA’s Southern Africa region, where large HIV programs in Lesotho, Mozambique, South Africa, Zambia, and Zimbabwe contribute patient-level data for dozens of small rural clinics within their respective program, we followed the approach used in prior surveys,^60^ purposively sampling sites that participated in IeDEA’s 2020 and 2023 surveys to both mitigate the logistical challenges of implementing the survey within these programs and facilitate longitudinal analyses of service availability and clinic resources. If all previously-surveyed clinics had exited a program, eligible sites were selected via random sampling from a list of clinics that remained active. Purposive sampling of sites that participated in IeDEA’s 2020 and 2023 surveys was also used for clinics operating within large service networks in the United States.

#### Survey implementation

The finalized questionnaire and clinic-specific survey links for 235 eligible sites were shared with IeDEA’s seven regional coordinating centers on August 31, 2026, with a target date for survey completion of November 30, 2026. At each site, the unique survey link and a printable copy of the survey are being shared with the director or designated staff of the HIV clinic. Survey instructions encourage respondents to consult with colleagues in other departments (e.g., laboratory, pharmacy), as needed, to complete the survey. To minimize the potential for social desirability bias, instructions also assure respondents of the confidentiality of their individual responses to the survey and encourage them to provide honest feedback on their day-to-day service delivery and routine practices, without concern that the data will be used to evaluate staff or clinic performance.

Clinics that completed the pilot survey are being asked only to respond to survey questions that were modified after the pilot. For clinics unable to complete an online questionnaire, responses marked on the printed questionnaire will be entered into REDCap by an IeDEA country or regional coordinator. Weekly updates on survey completion will be shared with IeDEA’s regional coordinating centers, and reminders will be sent to non-responding clinics to encourage participation. Data quality checks will be performed as survey responses are submitted, with all data checks and corrections completed by December 31, 2026, so that data can be released for planned analyses by early 2027.

### Data Harmonization

All survey data will be stored in a REDCap database hosted at VUMC. Survey data will be linked with data on clinic characteristics (e.g., facility type or level, urban vs. rural location) obtained from IeDEA’s central database of participating clinics. To complement data collected via the survey and enable concept leads to characterize the patient populations served by sites responding to the survey, patient data will be requested from IeDEA’s regional data managers. Summary metrics will be derived for the number of active patients served by responding clinics and programs, along with the distribution by sex at birth (male vs. female) and the broad age groups specified in investigator-linked research proposals (e.g., children 0-9 years, adolescents 10-19 years, young adults 20-24 years, adults 25+ years, and adults 40+ years). Active patients will be defined as the number of unique patients with at least one clinic visit or encounter (e.g., visit date, viral load test, or ART medication start date) during a calendar year. As database closure dates vary across IeDEA regions and cohorts, the most recent calendar year with complete data for each site will be used to derive summary metrics.

### Data analysis

Planned analyses will focus on 11 research topics, including analyses exploring the implementation status of new services and service modalities across IeDEA in 2026 and longitudinal analyses focused on changes in service capacity and availability between 2023 and 2026 (Table 1).

**Table 1.**
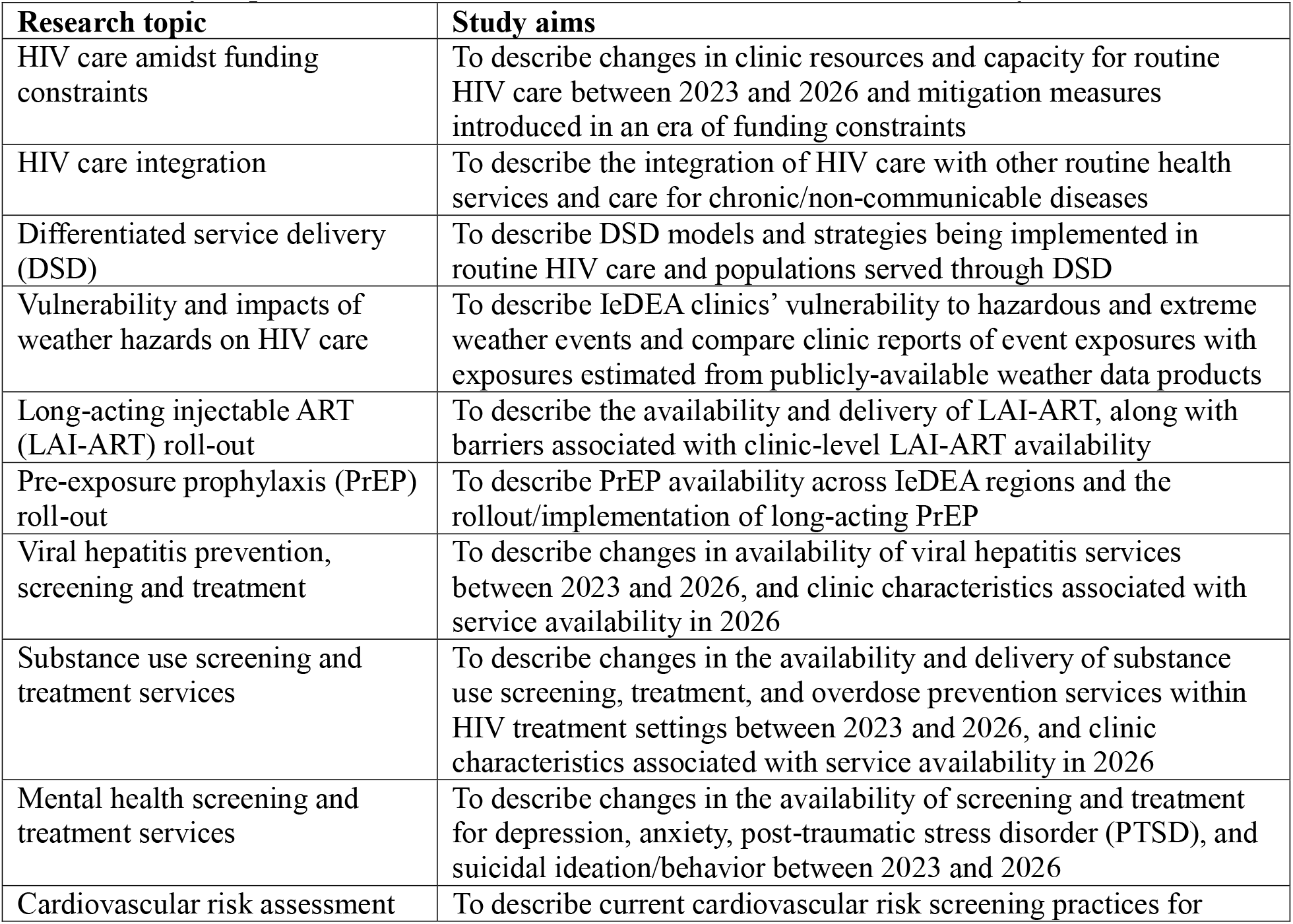
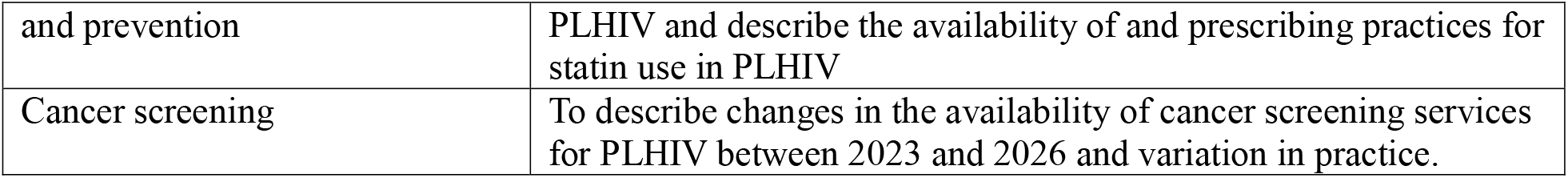
Analyses planned for IeDEA’s consortium-wide site assessment survey data.

### Study strengths and limitations

Strengths of IeDEA’s consortium-wide site assessment surveys include the diverse global cohort of participating HIV clinics across different levels of national health systems in low-, middle- and high-income countries. As IeDEA does not provide clinical or programmatic guidance to participating sites, IeDEA’s consortium-wide surveys provide a window into resources and HIV-related service delivery in real-world settings operating under national treatment guidelines. With response rates above 90% in prior IeDEA-wide surveys,^27 28 30 60^ the data are representative of sites participating in the consortium, and the mix of legacy survey content with new content on timely research questions proposed by IeDEA investigators will allow for the examination of temporal trends and the evolution of care as treatment guidelines and service delivery strategies change.

A limitation of IeDEA’s site assessment surveys is their reliance on voluntary self-report by HIV clinic directors or their designated staff. Knowledge about some specialized areas of care explored via our survey and care provided outside the HIV clinic may vary across clinics and respondents. Additionally, social desirability bias may induce some respondents to report practices aligned with local guidelines, even if their day-to-day practices differ because of resource constraints or other factors. Recall bias is an additional concern, particularly at sites where recent shifts in donor funding for the HIV response have resulted in staff lay-offs and/or reorganization of HIV care.^46 68-70^ Finally, sites participating in a global research consortium may not be representative of HIV service delivery within some countries and regions—particularly in contexts where IeDEA sites serve a sizable patient population and/or offer advanced levels of care.

## III. Ethics and dissemination

### Patient and public involvement

Key research topics of this study were informed by the expertise of a diverse group of investigators contributing to IeDEA’s scientific working groups and regional cohorts, as well as literature reviews performed during the preparation of research proposals for the survey. No patients were involved in the study’s design or survey development. Clinics participating in the pilot survey had the opportunity to comment on the survey content and provide feedback and suggestions for strengthening the questionnaire.

### Ethical considerations

The 2026 IeDEA-wide site assessment survey was reviewed by the VUMC Human Research Protections Program and given a non-research status per 45 CFR §46.102(l) (IRB#261052).

### Dissemination

Findings from planned analyses of the site assessment survey data will be shared with partners and stakeholders involved in the global HIV response, presented at regional and global scientific conferences focused on HIV-related care and published in peer-reviewed journals for a wide audience of clinicians and researchers. Relevant findings will also be shared with local partners within each IeDEA region. Additionally, in accordance with IeDEA’s standard operating procedures for multiregional research,^2^ site assessment survey data may be made available to other investigators to inform future implementation research or linked with patient-level data for approved implementation-observational hybrid studies^71^ that can inform HIV service delivery strategies.

## IV. Conclusion

This study will provide contemporary snapshots of the availability of routine HIV care and specialized services for PLWH in a global observational cohort of HIV clinics participating in the IeDEA consortium at a critical juncture. It will further document temporal trends in HIV service delivery and screening and management of infectious and non-infectious comorbidities that impact PLHIV, as well as real-world adoption and implementation of treatment guidelines and evidence-based practices in HIV care and historical hazardous weather exposure and impacts across diverse geographic and system settings.

Furthermore, it will contribute to existing assessments of service availability and practices, supporting a timely scientific agenda to inform policy and programs for improving HIV care outcomes, along with future research and analyses of IeDEA’s longitudinal patient data.

## Data Availability

All data produced in the present study will be available upon reasonable request to the authors

## Authors’ contributions

Contributors EB, FM, DN and SND, KW-K and AF designed this study. EB, FM, SA, MB, NVFV, MHK, KL, SL, AMP, RAP, AFRM, JeR, JoR and AS led the development and refinement of survey content. SND, DN, KW-K, AF and BM provided input on study procedures and data collection tools. FM developed the REDCap database for data collection. EB drafted the manuscript. All authors (EB, FM, SND, DN, KW-K, AF, BM, SA, MB, NVFV, MHK, KL, SL, AMP, RAP, AFRM, JeR, JoR and AS) participated in manuscript revisions. SA and EB created visualizations. All authors have read and approved the final manuscript.

## Acknowledgements

The authors thank the investigators, clinicians and data managers who contribute to IeDEA and participate in IeDEA’s site surveys, as well as members of the IeDEA Site Assessment Working Group.

