## Supplementary material 1 for "Documenting HIV service delivery and adoption of evidence-based practices in HIV care programs: A protocol for a cross-sectional survey of HIV clinics participating in the International epidemiology Databases to Evaluate AIDS (IeDEA)" (from another version of this preprint: version 1, 2026-09-17)

### IeDEA 2026 Site Assessment Survey

The **International Epidemiology Databases to Evaluate AIDS (IeDEA)** is conducting this survey among all health facilities that participate in IeDEA's research activities. IeDEA conducts this survey every 3 years to be able to describe HIV-related care and support at clinics participating in IeDEA. This survey covers **facility resources**, such as staffing, laboratory capacity, and medication availability, along with **routine HIV-related services** provided to patients enrolled in HIV care. The survey also includes sections on **special topics**, such as HIV prevention; screening and care for co-morbidities (e.g., hepatitis, cardiovascular disease, mental health, substance use, and cancers); the integration of HIV care with other health services; and the extent to which clinics participating in IeDEA experience extreme weather hazards.

This **survey is intended to be completed by the head of the HIV clinic, or other staff who have in-depth knowledge about the care and services provided in the HIV clinic and elsewhere within the health facility for people living with or at risk of HIV**. Most questions refer to care and services provided within the HIV clinic for adults living with HIV. A few questions in this survey may require consultation with staff in other units, such as laboratory and pharmacy departments. **If your health facility does not have a dedicated clinic for HIV care and treatment, please answer for the facility overall, regardless of what unit(s) serves patients with HIV. If your health facility has multiple HIV care and treatment clinics that serve different age-groups, please report on the services provided for adult patients living with HIV, unless otherwise indicated**. For clinics serving pediatric patients only, please report on the care you provide for pediatric patients.

We expect that the survey will take **approximately 1-2 hours to complete**. Remember that there are no incorrect answers to this survey. Your honest feedback on day-to-day service delivery and routine practices is important for understanding patient-level outcomes. Your answers to this survey will not be linked to you individually or used to evaluate staff or clinic performance in any way. Thank you for your time completing this survey. We are very grateful for your participation.

| | QUESTIONS | RESPONSES |
|---|---|---|
| 1 | **RESPONDENT INFORMATION** | |
| 1.1 | **Name of person completing this survey** | |
| 1.2 | **Email address of the person completing the survey** | |
| 1.3 | **Please enter the date this survey is being completed** | __ __/__ __/ 2026 (DD / MM / YYYY) |
| 1.4 | **What is your title?** | ☐ Head Clinician/Clinical Officer In-Charge<br>☐ Other clinician<br>☐ Site Manager<br>☐ Site Data Manager<br>☐ Head Nurse<br>☐ Other (specify) ________________________ |
| 2 | **CLINIC CHARACTERISTICS AND PATIENT POPULATION** | |
| 2.1 | **Is this clinic a public sector, non-governmental or faith-based organization, or a private/for-profit facility?** *Select one response only.* | ☐ Government/public<br>☐ Non-governmental or faith-based organization<br>☐ Private (for profit) |
| 2.2 | **How would you describe the area of residence of the population served by this health facility's HIV clinic(s)?** *Select one response only.* | ☐ Predominantly urban<br>☐ Predominantly rural<br>☐ Mixed urban/rural |
| 2.3 | **What types of patients with HIV are served at the HIV clinic(s)?** *Select all that apply. If your clinic serves any children or any adolescents within the age ranges listed, you may select that group, even if you do not serve all ages within the range specified.* | ☐ Children (ages 0-9)<br>☐ Adolescents (ages 10-19)<br>☐ Adults – general population (ages 20+) |
| 2.4 | **Does this health facility provide outpatient care only, or both outpatient and inpatient care?** *Select one response only.* | ☐ Outpatient care only<br>☐ Both outpatient and inpatient care |
| 2.5 | **Does your HIV clinic offer <u>dedicated</u> service hours or service days for any of the following patient groups?** *Dedicated means special hours or days when services are provided for a particular type of patient or population group. Please indicate which of the following groups are served through dedicated hours or days. Select "Not applicable" for any group not served by your clinic.* | |
| a | Infants and children (0−9 years) | ☐ Yes ☐ No ☐ Not applicable |
| b | Adolescents (10−19 years) | ☐ Yes ☐ No ☐ Not applicable |
| c | Pregnant and breastfeeding women | ☐ Yes ☐ No ☐ Not applicable |
| d | Adults (≥20 years) | ☐ Yes ☐ No ☐ Not applicable |
| e | At-risk populations (sex workers, men who have sex with men, people who inject drugs, etc.) | ☐ Yes ☐ No ☐ Not applicable |
| 2.6 | **Does your HIV clinic regularly offer patient services during any of the following times or days?** | |
| a | Early morning or evening hours | ☐ Yes ☐ No |
| b | Weekends | ☐ Yes ☐ No |
| c | Public holidays | ☐ Yes ☐ No |

| 3 | **CLINIC STAFFING** | | | |
|---|---|---|---|---|
| 3.1 | **How often are the following categories of staff available at this HIV clinic?** *Select one best response.* | **Available every day clinic is open** | **Available some days** | **Never available** |
| a | Pediatrician (generalist) | ☐ | ☐ | ☐ |
| b | Internist, family practitioner, generalist (physician) | ☐ | ☐ | ☐ |
| c | Infectious disease or HIV specialist | ☐ | ☐ | ☐ |
| d | Mid-level providers (clinical officers, nurses/nurse practitioners, midwives, physician assistants) | ☐ | ☐ | ☐ |
| e | Adherence counselors | ☐ | ☐ | ☐ |
| f | Peer educators/mentors/navigators | ☐ | ☐ | ☐ |
| g | Outreach workers | ☐ | ☐ | ☐ |
| h | Psychologist | ☐ | ☐ | ☐ |
| i | Psychiatrist | ☐ | ☐ | ☐ |

| 4 | **HIV TESTING & LABORATORY DIAGNOSTICS** | |
|---|---|---|
| 4.1 | **Which of the following types of <u>HIV testing services</u> are routinely offered by this health facility?** | |
| a | Facility-based testing (in the HIV clinic or elsewhere in the health facility) | ☐ Yes ☐ No ☐ Don't know |
| b | Community-based testing (e.g. HIV testing at workplaces and other community settings) | ☐ Yes ☐ No ☐ Don't know |
| c | HIV self-testing (i.e., any HIV self-testing kits made available by the facility for clients to collect their own samples and perform their test) | ☐ Yes ☐ No ☐ Don't know |
| d | Rapid HIV testing | ☐ Yes ☐ No ☐ Don't know |
| e | Partners/couples and/or index case testing | ☐ Yes ☐ No ☐ Don't know |
| f | "Family tree" testing (testing of family and other household members) | ☐ Yes ☐ No ☐ Don't know |

| 4.2 | **Where are the following <u>screening and diagnostic tests</u> performed for patients at this HIV clinic?** *Indicate whether diagnostic services are provided on-site (i.e., in the HIV clinic or elsewhere in the health facility), off-site via referral, or are not available for routine patient care. Select one best response.* | **On-site (in the HIV clinic or elsewhere in same health facility)** | **Only offsite (referral)** | **Not available** |
|---|---|---|---|---|
| a | DNA or RNA PCR for early infant diagnosis (EID) | ☐ | ☐ | ☐ |
| b | Quantitative PCR or HIV viral load testing | ☐ | ☐ | ☐ |
| c | HIV-1 genotypic drug resistance testing | ☐ | ☐ | ☐ |
| d | CD4 cell count testing | ☐ | ☐ | ☐ |
| e | Serum creatinine testing | ☐ | ☐ | ☐ |
| f | Cryptococcal meningitis screening (serum cryptococcal antigen or lateral flow assay) | ☐ | ☐ | ☐ |
| g | Cryptococcal meningitis diagnosis by CSF India Ink or latex agglutination | ☐ | ☐ | ☐ |
| h | Syphilis testing (RPR/TPHA/VDRL) | ☐ | ☐ | ☐ |

| 4.3 | **How often are the following laboratory and diagnostic tests offered at this facility?** *Select one best response. For tests only provided offsite, select "Test not available onsite."* | **Service available every day clinic is open** | **Service available some days** | **Test not available onsite** |
|---|---|---|---|---|
| a | HIV testing | ☐ | ☐ | ☐ |
| b | DNA or RNA PCR for early infant diagnosis (EID) | ☐ | ☐ | ☐ |
| c | Quantitative PCR or HIV viral load testing | ☐ | ☐ | ☐ |
| d | HIV-1 genotypic drug resistance testing | ☐ | ☐ | ☐ |
| e | CD4 cell count testing | ☐ | ☐ | ☐ |

| 4.4 | **For each of the following tests, what is the usual <u>turnaround time</u> for getting test results.** *<u>Turnaround time</u> means the time from ordering or referring a patient for the test to the time when results are received by the facility/clinic staff. Select one best response.* | **Turnaround time** | | | | | |
|---|---|---|---|---|---|---|---|
| | | **Same day (0 days)** | **1-7 days** | **8-14 days** | **15-30 days** | **> 30 days** | **Test not available** |
| a | HIV testing | ☐ | ☐ | ☐ | ☐ | ☐ | ☐ |
| b | DNA or RNA PCR for early infant diagnosis (EID) | ☐ | ☐ | ☐ | ☐ | ☐ | ☐ |
| c | Quantitative PCR or HIV viral load testing | ☐ | ☐ | ☐ | ☐ | ☐ | ☐ |
| d | HIV-1 genotypic drug resistance testing | ☐ | ☐ | ☐ | ☐ | ☐ | ☐ |
| e | CD4 cell count testing | ☐ | ☐ | ☐ | ☐ | ☐ | ☐ |

| 4.5 | **In the past 12 months, did this HIV clinic experience interruptions in routine viral load testing**? | ☐ Yes<br>☐ No **{SKIP to Q5.1}**<br>☐ Do not know **{SKIP to Q5.1}** |
|---|---|---|

| | | |
|---|---|---|
| 4.6 | **Which patients were prioritized when there were interruptions in routine viral load testing?**<br>*Select all that apply or "None".* | ☐ None (*No prioritization based on patient characteristics*)<br>☐ Pregnant/breastfeeding patients<br>☐ Infants, children or adolescents (<20 years)<br>☐ Patients returning to care after treatment interruption<br>☐ Patients with suspected treatment failure<br>☐ Patients with advanced HIV disease<br>☐ At-risk populations (people who inject drugs, sex workers, men who have sex with men, others) |

| | | |
|---|---|---|
| **5** | **CARE FOR PATIENTS NEWLY TESTING POSITIVE, RE-ENTERING CARE OR TRANSFERING FROM OTHER SITES** | |
| 5.1 | **How soon after confirming HIV diagnoses and/or treatment eligibility do patients routinely initiate ART?** *Routinely refers to the standard of care.*<br>*Select one best response.* | ☐ Same day that ART eligibility is established<br>☐ 1-7 days after establishing ART eligibility<br>☐ 8-14 days after establishing ART eligibility<br>☐ 15-30 days after establishing ART eligibility<br>☐ >30 days after establishing ART eligibility |
| 5.2 | **At this clinic, how many ART readiness counseling sessions are routinely conducted before eligible patients initiate ART?** *Routinely refers to the standard of care.*<br>*Select one best response.* | ☐ 0 sessions<br>☐ 1 session<br>☐ 2 sessions<br>☐ 3 sessions<br>☐ 4 or more sessions |
| 5.3 | **What types of support services are routinely provided at this facility to patients who receive a positive HIV test result?** *Routinely means provided as the standard of care.*<br>*Select all that apply or "None".* | ☐ None<br>☐ Psychosocial support from nurse, social worker, counselor, mentor, etc.<br>☐ Partner disclosure counseling and support<br>☐ Referral to support groups<br>☐ Referral to community-based volunteers/workers |
| 5.4 | **Is CD4 cell count testing done as the standard of care prior to ART initiation for newly-enrolling patients with HIV?** | ☐ Yes<br>☐ No |
| 5.5 | **Is CD4 cell count testing done as the standard of care prior to re-starting ART for patients re-entering care at this health facility?** | ☐ Yes<br>☐ No |
| 5.6 | **Which of the following screenings are routinely done at the time of enrollment into HIV care at this health facility (e.g., newly-diagnosed patients or patients who transfer to this site for HIV care)?** *Routinely means provided as the standard of care at enrollment.*<br>*Select all that apply or "None".* | ☐ None<br>☐ Pregnancy/breastfeeding<br>☐ Latent tuberculosis infection (LTBI)<br>☐ Tuberculosis (TB) disease<br>☐ Cryptococcal meningitis<br>☐ Sexually transmitted infections (STIs)<br>☐ Substance use disorders (alcohol, tobacco, or illicit drug use)<br>☐ Mental health disorders (depression, anxiety, post-traumatic stress, etc.) |

| | | | |
|---|---|---|---|
| **5.7** | **Which of the following ART initiation models are routinely offered by the staff of this HIV clinic?** | | |
| a | Facility-based ART initiation | ☐ Yes | ☐ No |
| b | Community ART initiation (mobile clinics in rural communities provide on-site ART initiation, community outreach events, initiation during community HIV campaigns) | ☐ Yes | ☐ No |
| c | Home-based ART initiation | ☐ Yes | ☐ No |

| | | | | | |
|---|---|---|---|---|---|
| **6** | **ART MONITORING, ADHERENCE & RETENTION STRATEGIES** | | | | |
| **6.1** | **What is the visit frequency for the following types of patients:** | **Every 1-2 months** | **Every 3-4 months** | **Every 5-6 months** | **>6 months** |
| a | Patients who are newly initiating ART | ☐ | ☐ | ☐ | ☐ |
| b | Patients who are stable on ART (on ART for at least 1 year and evidence of treatment success) | ☐ | ☐ | ☐ | ☐ |
| **6.2** | **What is the frequency of ART refills for the following types of patients:** | **Every 1-2 months** | **Every 3-4 months** | **Every 5-6 months** | **>6 months** |
| a | Patients who are newly initiating ART | ☐ | ☐ | ☐ | ☐ |
| b | Patients who are stable on ART (on ART for at least 1 year and evidence of treatment success) | ☐ | ☐ | ☐ | ☐ |
| c | Patients with advanced HIV disease (CD4 <200 cells/mm3 and/or WHO Clinical Stage 4 disease) | ☐ | ☐ | ☐ | ☐ |
| d | Patients on ART with virologic or therapeutic failure ("unstable clients" on ART >1 year) | ☐ | ☐ | ☐ | ☐ |

| | | | |
|---|---|---|---|
| **6.3** | **Which of the following treatment delivery models are routinely available at your HIV clinic?** | | |
| a | Individual models based at facility (fast track or quick pharmacy refill without clinical consultation) | ☐ Yes | ☐ No |
| b | Individual models not based at facility (mobile outreach, fixed community ART distribution points, home delivery). | ☐ Yes | ☐ No |
| c | Group models managed by health-care workers (provider-led treatment refill or support groups) | ☐ Yes | ☐ No |
| d | Group models managed by patients/clients (community ART refill group, patient-led community ART delivery) | ☐ Yes | ☐ No |

| | | |
|---|---|---|
| **6.4** | **How is ART medication adherence routinely monitored or assessed in patients at this HIV clinic?** *Routinely means that it is done as the standard of care.*<br><br>*Select all that apply or "Not applicable".* | ☐ Not applicable *(medication adherence not routinely monitored)*<br>☐ Patient self-report of medication adherence<br>☐ Review of medication pick-up/pharmacy refills<br>☐ Pill counts (Pharmacist/provider count of unused medication in pill bottles)<br>☐ Electronic dose monitoring (e.g., MEMS caps)<br>☐ Routine viral loads |

| | | |
|---|---|---|
| **6.5** | **Which of the following ART adherence support services are provided at this HIV clinic and which types of patients are offered these services?** *Select "None" if not available/not provided as part of adherence support, "All patients" if the adherence support is provided as standard of care for all patients, or "Eligible patients" if the adherence support is provided only to patients eligible for intensified or enhanced adherence support.* | ***Select one best response*** |
| a | One-on-one adherence counseling by HIV care providers or pharmacy staff | ☐ None ☐ All patients ☐ Eligible patients |
| b | Group adherence counseling by HIV care providers or pharmacy staff | ☐ None ☐ All patients ☐ Eligible patients |
| c | Facility-based ART adherence clubs | ☐ None ☐ All patients ☐ Eligible patients |
| d | Community-based ART adherence clubs | ☐ None ☐ All patients ☐ Eligible patients |
| e | Cash incentives | ☐ None ☐ All patients ☐ Eligible patients |
| f | Food or vitamin/micronutrient supplements | ☐ None ☐ All patients ☐ Eligible patients |
| g | Directly administered antiretroviral therapy (e.g., long-acting injectable ART) or directly observed treatment (DOT) | ☐ None ☐ All patients ☐ Eligible patients |

| | | |
|---|---|---|
| **6.6** | **Does this HIV clinic utilize text or voice messaging reminders to support any of the following:**<br>*Select all that apply or "None".* | ☐ None/Not applicable<br>☐ Medication adherence reminders<br>☐ Appointment reminders<br>☐ Follow-up of missed appointments |
| **6.7** | **What is done to follow-up with ART patients who miss appointments?**<br><br>*Select all that apply or "Nothing".* | ☐ Nothing (*no follow-up with ART patients who miss appointments*)<br>☐ Phone call to individual or family<br>☐ Message via online patient portal, SMS/WhatsApp, email or letter<br>☐ Home visit by clinic staff<br>☐ Home visit by community outreach worker |

| | | | |
|---|---|---|---|
| **6.8** | **Which of the following community-based activities does this HIV clinic or other partners currently support in the clinic catchment area?**<br>*For activities not currently supported in the clinic catchment area, please indicate whether they were previously supported by the clinic or other partners (e.g., health department or community-based organizations) in past 1-2 years.* | **Is community-based activity currently supported?** | **If "No", was this activity supported in the past 1-2 years by the clinic or other partners?** |
| a | ARV distribution | ☐ Yes ☐ No (→) | ☐ Yes ☐ No |
| b | Tracing patients lost to follow-up | ☐ Yes ☐ No (→) | ☐ Yes ☐ No |
| c | HIV prevention services and programs | ☐ Yes ☐ No (→) | ☐ Yes ☐ No |

| | | | | | |
|---|---|---|---|---|---|
| **6.9** | **Do patients with HIV routinely pay any fees (other than insurance co-pays) for the following types of routine and specialized services?**<br>*Select one best response, or "Not applicable" for any service that is not provided at this health facility* | **Yes** | **No** | **Don't know** | **Not applicable** |
| a | Routine clinic visits or consultations | ☐ | ☐ | ☐ | ☐ |
| b | Specialty clinic visits or consultations | ☐ | ☐ | ☐ | ☐ |
| c | ARVs | ☐ | ☐ | ☐ | ☐ |
| d | HIV testing | ☐ | ☐ | ☐ | ☐ |
| e | CD4 testing | ☐ | ☐ | ☐ | ☐ |
| f | Early infant diagnosis (EID) | ☐ | ☐ | ☐ | ☐ |
| g | HIV viral load testing | ☐ | ☐ | ☐ | ☐ |
| h | HIV drug resistance testing | ☐ | ☐ | ☐ | ☐ |

| | | |
|---|---|---|
| **6.10** | **In the past 12 months, did this HIV clinic have patients on a waiting list to receive ART?** | ☐ Yes<br>☐ No **{SKIP TO Q7.1}**<br>☐ Do not know **{SKIP TO Q7.1}** |
| **6.11** | **Which patients were prioritized for ART when the clinic had a waiting list?**<br><br>*Select all that apply or "None"* | ☐ None (*No prioritization based on patient characteristics*)<br>☐ Newly-enrolling/ART-naïve patients<br>☐ Patients already on treatment<br>☐ Patients with severe immunosuppression or comorbidities<br>☐ Pregnant/breastfeeding patients<br>☐ Infants and children (<10 years)<br>☐ Adolescents (10-19 years)<br>☐ At-risk populations (people who inject drugs, sex workers, men who have sex with men, others) |

| 7 | SERVICES PROVIDED TO PEDIATRIC AND ADOLESCENT PATIENTS | |
|---|---|---|
| 7.1 | **Which of the following services are provided at this health facility to <u>pediatric patients</u> (<10 years), including HIV-exposed infants?**<br><br>*Select all that apply or "Not Applicable" if no pediatric patients are served at this health facility.* | ☐ Not applicable *(no pediatric patients served at this facility)* **{→SKIP TO 7.3}**<br>☐ Postnatal ARV prophylaxis (prevention of transmission) to HIV-exposed infants<br>☐ ART initiation<br>☐ Infant feeding counseling<br>☐ Male circumcision for infants<br>☐ Immunizations<br>☐ Nutritional support<br>☐ Growth monitoring<br>☐ Integrated Management of Childhood Illness (IMCI)<br>☐ Screening for tuberculosis (TB) disease<br>☐ Testing for latent tuberculosis infection (LTBI) |
| 7.2 | **Does this health facility provide HIV care and treatment to infants <24 months of age?** | ☐ Yes<br>☐ No |
| 7.3 | **Does this health facility offer any of the following services for youth/adolescent patients with HIV?**<br><br>*Select all that apply or "None".* | ☐ None *(no dedicated services for youth/adolescent patients)*<br>☐ Dedicated hours or space for youth/adolescent HIV testing & counseling services<br>☐ Dedicated hours or space for youth/adolescent HIV care and treatment services<br>☐ Dedicated clinics or services during school holidays or weekends<br>☐ Peer counseling for youth/adolescent patients with HIV<br>☐ Support groups specifically for youth/adolescent patients with HIV<br>☐ Status disclosure counseling for youth/adolescents with perinatal HIV<br>☐ Services to support transition to adult HIV care |

| 8 | PHARMACY | | |
|---|---|---|---|
| 8.1 | **For each of the following medications, please indicate whether it is <u>dispensed/available at this health facility.</u>** For all medications <u>dispensed</u>, please indicate whether there were <u>supply disruptions/ stock-outs</u> lasting at least 1 week during the <u>past 12 months.</u> | **Medication dispensed/available** | **Stock-out lasting at least 1 week in past 12 months.** Select N/A if not dispensed |
| a | First-line HIV antiretroviral medications (ARVs) | ☐ Yes ☐ No ☐ Don't know | ☐ Yes ☐ No ☐ N/A |
| b | Second-line HIV ARVs | ☐ Yes ☐ No ☐ Don't know | ☐ Yes ☐ No ☐ N/A |
| c | Third-line HIV ARVs | ☐ Yes ☐ No ☐ Don't know | ☐ Yes ☐ No ☐ N/A |
| d | Long-acting injectable ARVs (combined cabotegravir/rilpivirine or injectable lenacapavir) | ☐ Yes ☐ No ☐ Don't know | ☐ Yes ☐ No ☐ N/A |
| e | Neonatal ARVs - Single-drug six-week prophylaxis for infants not at high risk of HIV acquisition (e.g., NVP, DTG or 3TC) | ☐ Yes ☐ No ☐ Don't know | ☐ Yes ☐ No ☐ N/A |
| f | Neonatal ARVs – Double or triple-drug regimen for infants at high risk of HIV acquisition (e.g., ABC/3TC-DTG). | ☐ Yes ☐ No ☐ Don't know | ☐ Yes ☐ No ☐ N/A |
| g | Dispersible (pediatric) dolutegravir | ☐ Yes ☐ No ☐ Don't know | ☐ Yes ☐ No ☐ N/A |
| h | TB prevention medications (INH, RIF) | ☐ Yes ☐ No ☐ Don't know | ☐ Yes ☐ No ☐ N/A |
| i | Drug-sensitive TB treatment medications (RHZE) | ☐ Yes ☐ No ☐ Don't know | ☐ Yes ☐ No ☐ N/A |
| j | Multidrug-resistant (MDR) TB treatment medications | ☐ Yes ☐ No ☐ Don't know | ☐ Yes ☐ No ☐ N/A |
| k | Cotrimoxazole (Bactrim, Septra, TMP-SMX) | ☐ Yes ☐ No ☐ Don't know | ☐ Yes ☐ No ☐ N/A |
| l | Fluconazole | ☐ Yes ☐ No ☐ Don't know | ☐ Yes ☐ No ☐ N/A |
| m | Amphotericin B | ☐ Yes ☐ No ☐ Don't know | ☐ Yes ☐ No ☐ N/A |
| n | Flucytosine (5FC) | ☐ Yes ☐ No ☐ Don't know | ☐ Yes ☐ No ☐ N/A |
| o | Short-acting contraceptives (pills, injectables, condoms) | ☐ Yes ☐ No ☐ Don't know | ☐ Yes ☐ No ☐ N/A |
| p | Long-acting contraceptives (implants, intrauterine devices) | ☐ Yes ☐ No ☐ Don't know | ☐ Yes ☐ No ☐ N/A |
| q | Selective serotonin reuptake inhibitors (SSRIs: Prozac, Zoloft, Paxil) | ☐ Yes ☐ No ☐ Don't know | ☐ Yes ☐ No ☐ N/A |
| r | Serotonin and norepinephrine reuptake inhibitors (SNRIs: Cymbalta, Effexor) | ☐ Yes ☐ No ☐ Don't know | ☐ Yes ☐ No ☐ N/A |
| s | Tricyclic antidepressants (amitriptyline, amoxapine, doxepin) | ☐ Yes ☐ No ☐ Don't know | ☐ Yes ☐ No ☐ N/A |
| t | Benzodiazepines (Xanax, Lorazepam, Klonopin) | ☐ Yes ☐ No ☐ Don't know | ☐ Yes ☐ No ☐ N/A |
| u | Antipsychotic medications (Haloperidol, Chlorpromazine, Fluphenazine) | ☐ Yes ☐ No ☐ Don't know | ☐ Yes ☐ No ☐ N/A |
| v | Mood stabilizers (Carbamazepine, Lithium, Valproate) | ☐ Yes ☐ No ☐ Don't know | ☐ Yes ☐ No ☐ N/A |
| w | Statins (e.g., pitavastatin, pravastatin, rosuvastatin, atorvastatin) | ☐ Yes ☐ No ☐ Don't know | ☐ Yes ☐ No ☐ N/A |

| 9 | PREVENTIVE SERVICES AND HIV PREVENTION | | | |
|---|---|---|---|---|
| 9.1 | **Which of the following preventive services are available at this health facility and where are these services routinely provided?** *Select one best response.* | **On-site (in the HIV clinic or elsewhere in same health facility)** | **Only offsite (referral)** | **Not available** |
| a | Prevention of mother-to-child/vertical transmission (PMTCT/PVT) | ☐ | ☐ | ☐ |
| b | Condoms | ☐ | ☐ | ☐ |
| c | Voluntary male medical circumcision services | ☐ | ☐ | ☐ |
| d | Family planning/contraceptive methods other than condoms | ☐ | ☐ | ☐ |
| e | HPV vaccine | ☐ | ☐ | ☐ |

| | | |
|---|---|---|
| 9.2 | **Is pre-exposure prophylaxis (PrEP), an oral or injectable medication to prevent HIV infection provided at this health facility?** | ☐ Yes **{SKIP TO Q9.3}**<br>☐ No |
| a | **Are there any plans to provide PrEP services at this health facility (either in the HIV clinic or elsewhere in the health facility)?** | ☐ Yes<br>☐ No<br>☐ Don't know |
| b | **Where is PrEP available for patients/clients at this health facility?** | ☐ Offsite via referral to other health facility **{SKIP TO Q10.1}**<br>☐ Not available/no referrals for PrEP **{SKIP TO Q10.1}** |

| 9.3 | **Which of the following units/departments provide PrEP at this health facility?** | | | |
|---|---|---|---|---|
| a | HIV clinic | ☐ Yes | ☐ No | ☐ Don't know |
| b | General outpatient/primary health care unit | ☐ Yes | ☐ No | ☐ Don't know |
| c | Sexually transmitted infection unit | ☐ Yes | ☐ No | ☐ Don't know |
| d | Family planning unit | ☐ Yes | ☐ No | ☐ Don't know |
| e | Maternal health unit | ☐ Yes | ☐ No | ☐ Don't know |
| 9.4 | **Which of the following PrEP formulations for HIV prevention are provided at this health facility?** | | | |
| a | Oral pills (Emtricitabine/Tenofovir disoproxil fumarate (TDF) or lamivudine/TDF) | ☐ Yes | ☐ No | ☐ Don't know |
| b | Long-acting injectable cabotegravir (CAB-LA) | ☐ Yes | ☐ No | ☐ Don't know |
| c | Long-acting injectable lenacapavir (LEN) | ☐ Yes | ☐ No | ☐ Don't know |
| d | Dapivirine vaginal ring (DVR) | ☐ Yes | ☐ No | ☐ Don't know |

| 9.5 | **Which of the following population groups are currently receiving PrEP services at this health facility?** *Select "Not applicable" for any group that does not generally access/seek health care at this health facility.* | | | | |
|---|---|---|---|---|---|
| a | Pregnant and breastfeeding women | ☐ Yes | ☐ No | ☐ Don't know | ☐ Not applicable |
| b | Non-pregnant, not breastfeeding women | ☐ Yes | ☐ No | ☐ Don't know | ☐ Not applicable |
| c | Gay, bisexual and other men who have sex with men | ☐ Yes | ☐ No | ☐ Don't know | ☐ Not applicable |
| d | Female or male sex workers | ☐ Yes | ☐ No | ☐ Don't know | ☐ Not applicable |
| e | Heterosexual men | ☐ Yes | ☐ No | ☐ Don't know | ☐ Not applicable |
| f | People with serodiscordant or serodifferent partner(s) | ☐ Yes | ☐ No | ☐ Don't know | ☐ Not applicable |
| g | People who inject drugs | ☐ Yes | ☐ No | ☐ Don't know | ☐ Not applicable |
| h | People who are incarcerated or in other closed settings | ☐ Yes | ☐ No | ☐ Don't know | ☐ Not applicable |

| | | | | |
|---|---|---|---|---|
| 9.6 | **Do patients routinely pay any fees (other than insurance co-pays) for PrEP services?** | ☐ Yes | ☐ No | ☐ Don't know |

| 9.7 | **Which types of providers initiate/prescribe PrEP at this health facility?** *Select "not applicable" for providers not at your health facility* | | | | |
|---|---|---|---|---|---|
| a | Physician | ☐ Yes | ☐ No | ☐ Don't know | ☐ Not applicable |
| b | Mid-level providers (generalist clinical officers, nurses/nurse practitioners, physician assistants) | ☐ Yes | ☐ No | ☐ Don't know | ☐ Not applicable |
| c | Pharmacist | ☐ Yes | ☐ No | ☐ Don't know | ☐ Not applicable |
| d | Other | ☐ Yes | ☐ No | ☐ Don't know | ☐ Not applicable |

| 9.8 | **Which of the following resources for PrEP provision are available at this health facility?** | | | |
|---|---|---|---|---|
| a | Guidelines or policies on PrEP (who should receive it, how to prescribe it, etc.) | ☐ Yes | ☐ No | ☐ Don't know |
| b | Medical providers trained in PrEP | ☐ Yes | ☐ No | ☐ Don't know |
| c | Dedicated space for PrEP counseling and services | ☐ Yes | ☐ No | ☐ Don't know |
| d | Medical record system for tracking PrEP patients | ☐ Yes | ☐ No | ☐ Don't know |
| 9.9 | ***Optional*: What are the main challenges this health facility is currently facing in providing PrEP or retaining patients on PrEP?** | | | |

| 10 | LONG-ACTING INJECTABLE ANTIRETROVIRAL THERAPY (LAI-ART) | | | |
|---|---|---|---|---|
| 10.1 | **Which of the following types of LAI-ART are currently available at this HIV clinic?** *Select one best response for each type of LAI-ART, or "Not available". If LAI-ART is available, please indicate whether it is a routine service or provided through a pilot initiative or research study.* | **Available as routine service** | **Available through pilot or research initiative** | **Not available** |
| a | Long acting injectable cabotegravir/rilpivirine (LAI-CAB/RPV) | ☐ | ☐ | ☐ |
| b | Long acting injectable lenacapavir (LAI-LEN) | ☐ | ☐ | ☐ |

| | | |
|---|---|---|
| 10.2 | **Which of the following staff administer LAI-ART at your clinic?**<br><br>*Select all that apply or "Not applicable" if LAI-ART is not available.* | ☐ Not applicable (LAI-ART not available)<br>☐ Physicians<br>☐ Nurses/nurse practitioners<br>☐ Pharmacists<br>☐ Physician assistants<br>☐ Lay providers/personnel (trained non-health professionals)<br>☐ Other |
| 10.3 | **Which of the following are challenges in providing LAI-ART at this clinic?**<br><br>*If your clinic does not currently provide LAI-ART please select all the reasons why LAI-ART is not provided. Multiple reasons may be selected.* | ☐ None (*no challenges providing LAI-ART*)<br>☐ Difficulty procuring LAI-ART medications<br>☐ Difficulty storing LAI-ART medications<br>☐ Lack of providers trained to administer LAI-ART<br>☐ Lack of private space for administration of LAI-ART<br>☐ Lack of supplies/commodities for administration of LAI-ART<br>☐ Lack of clinical guidelines on LAI-ART<br>☐ Patient record-keeping systems are not updated for recording LAI-ART<br>☐ Difficulty tracking LAI-ART appointments and follow-up<br>☐ Cost of LAI-ART medications to patients<br>☐ Patients' lack of awareness of LAI-ART<br>☐ Lack of patients clinically eligible for LAI-ART<br>☐ Don't know<br>☐ Other (specify)______________________________ |

| | | |
|---|---|---|
| **11** | **VIRAL HEPATITIS-RELATED PREVENTION, SCREENING, DIAGNOSIS & TREATMENT** | |
| **11.1** | **Which of the following hepatitis screenings are routinely performed at the time of enrollment into HIV care at this health facility (e.g., newly-diagnosed patients or patients who transfer to this site for HIV care)?** | |
| a | Hepatitis B virus (HBV) | ☐ Yes ☐ No |
| b | Hepatitis C virus (HCV) | ☐ Yes ☐ No |
| c | Hepatitis D virus (HDV) - Delta hepatitis | ☐ Yes ☐ No |
| **11.2** | **Which of the following hepatitis screenings are routinely performed during follow-up visits for enrolled patients with HIV?** | |
| a | New (incident) HBV | ☐ Yes ☐ No |
| b | New (incident) HCV | ☐ Yes ☐ No |
| c | New (incident) HDV | ☐ Yes ☐ No |

| | | | | |
|---|---|---|---|---|
| **11.3** | **Where are the following laboratory and diagnostic tests for hepatitis routinely performed for patients enrolled in care at this HIV clinic?** *Select one best response for each test listed. Select "Not available" for any test not available on-site or via referral.* | **On-site (in the HIV clinic or elsewhere in same health facility)** | **Only offsite (referral)** | **Not available** |
| a | Hepatitis B core antibody (anti-HBc) to measure past or active (chronic) HBV | ☐ | ☐ | ☐ |
| b | Hepatitis B surface antigen (HBsAg) to measure active (chronic) HBV | ☐ | ☐ | ☐ |
| c | Hepatitis B viral load (HBV DNA) to measure active (chronic) HBV or response to HBV antiviral therapy | ☐ | ☐ | ☐ |
| d | Hepatitis C antibody (anti-HCV) to measure past or active (chronic) HCV infection | ☐ | ☐ | ☐ |
| e | Hepatitis C viral load (HCV RNA) to measure active (chronic) HCV infection or response to HCV antiviral therapy | ☐ | ☐ | ☐ |
| f | Hepatitis D antibody (anti-HDV) to measure past or active (chronic) HDV infection | ☐ | ☐ | ☐ |
| g | Hepatitis D viral load (HDV RNA) to measure active (chronic) HDV infection or response to HDV antiviral therapy | ☐ | ☐ | ☐ |
| **11.4** | **Where are the following hepatitis prevention and treatment services provided to enrolled patients with HIV?** *Select one best response for each service listed. Select "Not available" for any service not available on-site or via referral.* | **On-site (in the HIV clinic or elsewhere in same health facility)** | **Only offsite (referral)** | **Not available** |
| a | HBV vaccine | ☐ | ☐ | ☐ |
| b | Treatment for HBV with antiviral medication | ☐ | ☐ | ☐ |
| c | Treatment for HCV with antiviral medication | ☐ | ☐ | ☐ |
| d | Treatment for HDV (Delta hepatitis) with antiviral medication | ☐ | ☐ | ☐ |

| | | |
|---|---|---|
| **12** | **SCREENING AND TREATMENT FOR SUBSTANCE USE DISORDERS FOR PATIENTS LIVING WITH HIV** | |
| **12.1** | **Which structured instrument(s) are used at this health facility to screen patients with HIV for alcohol use disorders?**<br><br>*Select all that apply or "Not applicable" if patients with HIV are not screened for alcohol use disorders.* | ☐ Screening is done without using any specific questionnaire or tool (for example, by asking patients directly or based on clinical judgment)<br>☐ Alcohol Use Disorders Identification Test (AUDIT) or AUDIT-C<br>☐ Alcohol, Smoking, and Substance Involvement Screening Test (ASSIST)<br>☐ Cut down, Annoyed, Guilty, Eye-opener (CAGE)<br>☐ Other (specify)______________________________<br>☐ Don't know<br>☐ Not applicable (no screening for alcohol use disorders) |

| | | |
|---|---|---|
| 12.2 | **Which types of treatment interventions for <u>alcohol use disorders</u> are available at this health facility for patients with HIV?**<br>*Select all that apply or "None."* | ☐ None *(no treatment available at this health facility)*<br>☐ Counseling or brief intervention (counseling/psychotherapy, SBIRT)<br>☐ Detoxification services or hospitalization<br>☐ Pharmacological treatment (e.g., disulfiram, naltrexone, acamprosate)<br>☐ Other (specify)________________________________________ |
| 12.3 | **At this health facility, which of the following <u>substance use disorders</u> (other than alcohol use) are patients with HIV screened for?** *<u>Screening</u> refers to any type of structured or unstructured assessment.*<br>*Select all that apply or "None" if screening for substance use disorders is not performed.* | ☐ None (no screening for substance use disorders) **{SKIP TO Q12.5}**<br>☐ Tobacco (e.g., smoked, smokeless)<br>☐ Cannabis (marijuana)<br>☐ Cocaine/crack<br>☐ Ecstasy and other club drugs<br>☐ Hallucinogens<br>☐ Methamphetamine<br>☐ Opioids<br>☐ Other |
| 12.4 | **Which structured instrument(s) are used to screen patients with HIV for <u>substance use disorders</u> (other than alcohol use)?**<br>*Select all that apply or "None."* | ☐ Screening is done without using any specific questionnaire or tool<br>☐ Addiction Severity Index (ASI)<br>☐ Alcohol, Smoking, and Substance Involvement Screening Test (ASSIST)<br>☐ Drug Abuse Screening Test (DAST)<br>☐ Other (specify)________________________________________<br>☐ Don't know |
| 12.5 | **Which types of treatment interventions for <u>substance use disorders</u> (other than alcohol use) are available at this health facility for patients with HIV?**<br>*Select all that apply or "None."* | ☐ None (*no treatment available at this health facility*)<br>☐ Counseling or brief intervention (counseling/psychotherapy, SBIRT)<br>☐ Detoxification services or hospitalization<br>☐ Medications for opioid use disorder (methadone, buprenorphine)<br>☐ Nicotine replacement<br>☐ Other pharmacological treatment<br>☐ Other (specify)________________________________________ |
| 12.6 | **Which harm reduction services are provided at this health facility?**<br>*Select all that apply, or "None".* | ☐ Clean needles or syringes<br>☐ Naloxone or overdose prevention education<br>☐ None<br>☐ Do not know |

| 12.7 | Where are the following services performed for patients with HIV at this health facility? | Provided in HIV Clinic | In same health facility (but not at HIV clinic) | Only offsite (referral) | Not available |
|---|---|---|---|---|---|
| a | Screening for substance use disorders (alcohol and/or drugs) | ☐ | ☐ | ☐ | ☐ |
| b | Counseling or brief intervention (counseling/psychotherapy, SBIRT) for substance use disorders (alcohol and/or drugs) | ☐ | ☐ | ☐ | ☐ |
| c | Pharmacological treatment or medication (naltrexone, acamprosate, disulfiram; methadone or buprenorphine) | ☐ | ☐ | ☐ | ☐ |
| d | Detoxification or hospitalization | ☐ | ☐ | ☐ | ☐ |

| 13 | SCREENING AND TREATMENT FOR MENTAL HEALTH DISORDERS FOR PATIENTS LIVING WITH HIV | | | | |
|---|---|---|---|---|---|
| 13.1 | **Which patients with HIV are routinely screened for the following disorders?** *Screening refers to any type of structured or unstructured assessment.* | **None (No patients routinely screened).** | **All patients** | **Patients with mental health symptoms** | **Patients with virologic failure or poor ART adherence** |
| a | Depression | ☐ | ☐ | ☐ | ☐ |
| b | Anxiety | ☐ | ☐ | ☐ | ☐ |
| c | Post-traumatic stress disorder (PTSD) | ☐ | ☐ | ☐ | ☐ |
| d | Suicidal ideation | ☐ | ☐ | ☐ | ☐ |

| | | |
|---|---|---|
| 13.2 | **Which structured instrument(s) are used at this health facility to screen patients with HIV for depression?**<br>*Select all that apply or "None." If patients with HIV are not routinely screened for depression, select "Not applicable."* | ☐ None (no structured or standardized depression screening tool used)<br>☐ Beck Depression Inventory (BDI)<br>☐ Center for Epidemiologic Studies Depression Scale (CES-D)<br>☐ Hamilton Rating Scale for Depression (HAM-D)<br>☐ Hospital Anxiety and Depression Scale (HAD)<br>☐ Patient Health Questionnaire (e.g., PHQ-2, PHQ-4, PHQ-9)<br>☐ Other (specify)________________________________________<br>☐ Do not know<br>☐ Not applicable (no patients routinely screened for depression) |

| | | |
|---|---|---|
| 13.3 | **Which structured instrument(s) are used at this health facility to screen patients with HIV for anxiety?** *Select all that apply or "None." If patients with HIV are not routinely screened for anxiety, select "Not applicable."* | ☐ None (no structured or standardized screening tool used)<br>☐ Beck Anxiety Inventory (BAI)<br>☐ Generalized Anxiety Disorder 7-item scale (GAD-7)<br>☐ Hospital Anxiety and Depression Scale (HAD)<br>☐ State-Trait Anxiety Inventory (STAI)<br>☐ Other (specify)__________________________<br>☐ Don't know<br>☐ Not applicable (no patients routinely screened for anxiety) |
| 13.4 | **Which structured instrument(s) are used at this health facility to screen patients with HIV for PTSD?** *Select all that apply or "None." If patients with HIV are not routinely screened for PTSD, select "Not applicable."* | ☐ None (no structured PTSD screening tool used)<br>☐ Life Event Checklist<br>☐ Primary Care PTSD Screen (PC-PTSD)<br>☐ PTSD Checklist for DSM-5 (PCL-5)<br>☐ Short PTSD Rating Interview (SPRINT)<br>☐ Trauma Screening Questionnaire (TSQ)<br>☐ Other (specify)__________________________<br>☐ Don't know<br>☐ Not applicable (no patients routinely screened for PTSD) |
| 13.5 | **Which structured instrument(s) are used at this health facility to screen patients with HIV for suicidal ideation or behavior?** *Select all that apply or "None". If patients with HIV are not routinely screened for suicidal ideation, select "Not applicable".* | ☐ None (no structured screening tool for suicidal ideation or behavior used)<br>☐ Columbia Suicide Severity Rating Scale (C-SSRS)<br>☐ Ask Suicide-Screener<br>☐ Other (specify)__________________________<br>☐ Don't know<br>☐ Not applicable (no patients routinely screened for suicidal ideation) |

| 13.6 | Which types of therapy or treatments are available at this health facility for patients with HIV with the following disorders? | Counseling or psychotherapy | Medication | **None** *(No therapy or treatment available at health facility)* | Don't know |
|---|---|---|---|---|---|
| a | Depression | ☐ | ☐ | | ☐ |
| b | Anxiety | ☐ | ☐ | | ☐ |
| c | PTSD | ☐ | ☐ | | ☐ |

| | | |
|---|---|---|
| 13.7 | **Which type of service provider typically provides mental health counseling or therapy for patients with HIV at your health facility?** *Select all that apply.* | ☐ Psychiatrists, psychologists, or other mental health professionals<br>☐ Generalist physicians, mid-level providers (clinical officers, nurses, midwives), or other health professionals without specialized mental health training<br>☐ Peers or other non-clinical staff<br>☐ Other<br>☐ Do not know<br>☐ Not applicable (mental health counseling/ therapy not provided at this facility) |
| 13.8 | **Is there a standard safety protocol at this health facility for responding to patients with suicidal ideation or behavior?** *A safety protocol refers to a standardized procedure that guides staff on how to respond to suicide risk.* | ☐ Yes<br>☐ No<br>☐ Do not know |
| 13.9 | **Is a safety plan developed at this health facility for patients with HIV at risk of suicide?** *A safety plan refers to a personalized plan to help a person recognize warning signs, use coping strategies, and access support during a suicidal crisis.* | ☐ Yes<br>☐ No<br>☐ Do not know |
| 13.10 | **Are emergency interventions (such as crisis services) available for patients with HIV at risk of suicide, when needed?** | ☐ Yes<br>☐ No<br>☐ Do not know |

| 14 | CARDIOVASCULAR RISK SCREENING AND MANAGEMENT | | | |
|---|---|---|---|---|
| 14.0 | **Do you serve patients with HIV who are aged 40 years or older?** | ☐ Yes ☐ No **{SKIP TO 15.0)** | | |
| 14.1 | **Where are the following evaluations performed for patients with HIV at this health facility?** *Select one best response or "Not available".* | **On-site (in the HIV clinic or elsewhere in same health facility)** | **Only offsite (referral)** | **Not available** |
| a | Weight | ☐ | ☐ | ☐ |
| b | Height | ☐ | ☐ | ☐ |
| c | Blood pressure | ☐ | ☐ | ☐ |
| d | Lipid profile (e.g, total cholesterol, LDL cholesterol, HDL cholesterol) | ☐ | ☐ | ☐ |
| e | Fasting glucose | ☐ | ☐ | ☐ |
| f | Hb1AC | ☐ | ☐ | ☐ |
| g | Electrocardiogram | ☐ | ☐ | ☐ |
| h | Echocardiogram | ☐ | ☐ | ☐ |

| | | |
|---|---|---|
| 14.2 | **At this health facility, which providers are responsible for evaluating risks for developing cardiovascular disease among patients with HIV?** *Select all that apply or "Not applicable" if cardiovascular risk evaluation is not performed.* | ☐ Internist, family practitioner, generalist (physician)<br>☐ Infectious disease or HIV specialist<br>☐ Mid-level provider (clinical officers, nurses/nurse practitioners, physician assistants)<br>☐ Cardiologist<br>☐ Other<br>☐ Not applicable **{SKIP TO Q14.6}** |
| 14.3 | **At this health facility, which patients with HIV are assessed for cardiovascular risk?** *Select all that apply.* | ☐ All patients<br>☐ Only patients ≥ 40 years old<br>☐ Only patients with comorbidities/risks (e.g., diabetes mellitus, hypertension, dyslipidemia, myocardial infarction, chronic renal disease, metabolic syndrome, active smoking)<br>☐ Don't know |
| 14.4 | **At this health facility, which cardiovascular risk scoring method(s) are used to estimate cardiovascular risk in patients with HIV?** *Select all that apply.* | ☐ Framingham score (FRS)<br>☐ Atherosclerotic Cardiovascular Disease (ASCVD) Risk Score (ACC/AHA)<br>☐ Systematic Coronary Risk Evaluation (SCORE2)<br>☐ Data Collection Adverse Events of Anti-HIV drugs (D:A:D)<br>☐ Pooled Cohort Equations/Predicting Risk of Cardiovascular Events (PCE/Prevent)<br>☐ WHO HEARTS risk score<br>☐ Other (specify)________________________<br>☐ Don't know |
| 14.5 | **At this health facility, how often is cardiovascular risk assessed in patients with HIV with no known cardiovascular disease?** *Select one best response.* | ☐ At entry to HIV care only<br>☐ At every visit<br>☐ Annually<br>☐ Every 2-3 years<br>☐ At the time of a cardiovascular event (myocardial infarction, stroke)<br>☐ Other<br>☐ Don't know |
| 14.6 | **At this health facility, are statins prescribed for primary prevention in HIV patients who have not had a prior cardiovascular event? (e.g, myocardial infarction or stroke)** | ☐ Yes (Statins prescribed for primary prevention)<br>☐ No (Statins only prescribed after a cardiovascular event) **{SKIP TO Q14.8}**<br>☐ Not applicable (Statins not routinely prescribed at this site) **{SKIP TO Q15.0}** |
| 14.7 | **At this HIV clinic, what guides decisions to prescribe statins for primary prevention in people living with HIV?** | ☐ Cardiovascular risk score<br>☐ Other assessments (not based on a formal risk score)<br>☐ Don't know |
| 14.8 | **Which of the following types of statins can you currently prescribe for patients living with HIV?** *Select all that apply or "Don't know".* | ☐ Atorvastatin ☐ Pravastatin ☐ Lovastatin ☐ Don't know<br>☐ Rosuvastatin ☐ Simvastatin ☐ Pitavastatin |

| 15 | **CANCER SCREENING PROVIDED TO PATIENTS WITH HIV** | | | | | |
|---|---|---|---|---|---|---|
| 15.0 | **Do you serve patients with HIV who are aged 25 years or older?** | ☐ Yes ☐ No **{SKIP TO 16.1)** | | | | |
| 15.1 | **Where are the following cancer screenings routinely performed for patients with HIV?** *Select one best response for each cancer screening or "Not available" for cancer screenings not performed. Select "Don't know" if you are unfamiliar with any cancer screening type listed.* | **Provided in HIV Clinic** | **In same health facility (but not at HIV clinic)** | **Only offsite (referral)** | **Not available** | **Don't know** |
| a | Cervical cancer screening by visual inspection (VIA or VILI) | ☐ | ☐ | ☐ | ☐ | ☐ |
| b | Cervical cancer screening by cytology (Pap smear) | ☐ | ☐ | ☐ | ☐ | ☐ |
| c | Molecular cervical HPV testing (self-collected or provider-collected) | ☐ | ☐ | ☐ | ☐ | ☐ |
| d | Anal cancer screening by cytology (anal Pap test) | ☐ | ☐ | ☐ | ☐ | ☐ |
| e | Molecular anal HPV testing | ☐ | ☐ | ☐ | ☐ | ☐ |
| f | High-resolution anoscopy (HRA) | ☐ | ☐ | ☐ | ☐ | ☐ |
| g | Anal cancer screening by Digital Anorectal Exam (DARE) | ☐ | ☐ | ☐ | ☐ | ☐ |
| h | Ultrasound for liver cancer screening | ☐ | ☐ | ☐ | ☐ | ☐ |
| i | Other liver cancer screening tests (computed tomography (CT) scan or serum alpha fetoprotein) | ☐ | ☐ | ☐ | ☐ | ☐ |
| j | Breast exam by provider | ☐ | ☐ | ☐ | ☐ | ☐ |
| k | Breast mammography or ultrasound | ☐ | ☐ | ☐ | ☐ | ☐ |
| l | Colon cancer screening by colonoscopy | ☐ | ☐ | ☐ | ☐ | ☐ |
| m | Colon cancer screening by fecal occult blood tests | ☐ | ☐ | ☐ | ☐ | ☐ |
| n | Lung cancer screening (by CT scan) | ☐ | ☐ | ☐ | ☐ | ☐ |
| o | Prostate cancer screening by prostate-specific antigen (PSA) laboratory test | ☐ | ☐ | ☐ | ☐ | ☐ |

| 15.2 | **What types of patients are routinely screened for the following cancers?** *Select one best response for each cancer screening type.* | **All patients (male and female) of eligible age (per clinical guidelines)** | **Male patients of eligible age (per clinical guidelines)** | **Female patients of eligible age (per clinical guidelines)** | **Only symptomatic patients** | **None / no patients routinely screened** |
|---|---|---|---|---|---|---|
| a | Cervical cancer | Not applicable | Not applicable | ☐ | ☐ | ☐ |
| b | Anal cancer | ☐ | ☐ | ☐ | ☐ | ☐ |
| c | Liver cancer | ☐ | ☐ | ☐ | ☐ | ☐ |
| d | Breast cancer | ☐ | ☐ | ☐ | ☐ | ☐ |
| e | Colon cancer | ☐ | ☐ | ☐ | ☐ | ☐ |
| f | Lung cancer | ☐ | ☐ | ☐ | ☐ | ☐ |
| g | Prostate cancer | Not applicable | ☐ | Not applicable | ☐ | ☐ |
| 15.3 | **What clinic barriers limit or impede offering routine screening for the following cancers to patients with HIV at this health facility?** *Multiple barriers may be selected. Select all that apply for each cancer screening type or "None".* | **Lack of trained staff to perform screening** | **Lack of equipment or supplies to perform screening** | **Lack of standardized or national screening guidelines to inform clinic policy** | **Inability to refer for specialist evaluation of abnormal results** | **None / no barriers for screening** |
| a | Cervical cancer | ☐ | ☐ | ☐ | ☐ | ☐ |
| b | Anal cancer | ☐ | ☐ | ☐ | ☐ | ☐ |
| c | Liver cancer | ☐ | ☐ | ☐ | ☐ | ☐ |
| d | Breast cancer | ☐ | ☐ | ☐ | ☐ | ☐ |
| e | Colon cancer | ☐ | ☐ | ☐ | ☐ | ☐ |
| f | Lung cancer | ☐ | ☐ | ☐ | ☐ | ☐ |
| g | Prostate cancer | ☐ | ☐ | ☐ | ☐ | ☐ |

| 16 | **INTEGRATED SERVICE DELIVERY FOR HIV AND OTHER CHRONIC CONDITIONS** | |
|---|---|---|
| 16.1 | **Which of the following types of patients receive care in the clinic/department that provides HIV care?** *Select all that apply* | ☐ People living with HIV<br>☐ Adults at risk or exposed to HIV<br>☐ Infants exposed to HIV<br>☐ Other people without HIV (apart from at-risk or exposed adults/infants) |
| 16.2 | **Does the HIV clinic/department provide any other health care services such as TB treatment, general outpatient services, or care for non-communicable diseases such as diabetes, hypertension, mental health, or substance use disorders?** | ☐ Yes<br>☐ No **{SKIP TO Q16.4}** |

| 16.3 | **For each of the following services, to what extent are all aspects of care provided in the HIV clinic and what is the extent of training received by providers in the HIV clinic?** | | |
|---|---|---|---|
| | **Service** | **Services provided in the HIV clinic?** | **Providers in HIV clinic trained?** |
| a | Tuberculosis (TB) care | ☐ All aspects of TB care<br>☐ Some aspects of TB care<br>☐ TB care not provided **{SKIP TO Q16.3b}** | ☐ All providers trained in TB care<br>☐ Some providers trained in TB care<br>☐ No providers trained in TB care |
| b | Sexual and reproductive health (SRH) care | ☐ All aspects of SRH care<br>☐ Some aspects of SRH care<br>☐ SRH care not provided **{SKIP TO Q16.3c}** | ☐ All providers trained in SRH care<br>☐ Some providers trained in SRH care<br>☐ No providers trained in SRH care |
| c | Hypertension (HTN) care | ☐ All aspects of HTN care<br>☐ Some aspects of HTN care<br>☐ HTN care not provided **{SKIP TO Q16.3d}** | ☐ All providers trained in HTN care<br>☐ Some providers trained in HTN care<br>☐ No providers trained in HTN care |
| d | Diabetes Mellitus (DM) care | ☐ All aspects of DM care<br>☐ Some aspects of DM care<br>☐ DM care not provided **{SKIP TO Q16.3e}** | ☐ All providers trained in DM care<br>☐ Some providers trained in DM care<br>☐ No providers trained in DM care |
| e | Mental health (MH) care - depression, anxiety, etc. | ☐ All aspects of MH care<br>☐ Some aspects of MH care<br>☐ MH care not provided **{SKIP TO Q16.3f}** | ☐ All providers trained in MH care<br>☐ Some providers trained in MH care<br>☐ No providers trained in MH care |
| f | Substance use (SU) care | ☐ All aspects of SU care provided in HIV clinic<br>☐ Some aspects of SU care provided in HIV clinic<br>☐ SU care not provided in HIV clinic **{SKIP TO Q16.4}** | ☐ All providers trained in SU care<br>☐ Some providers trained in SU care<br>☐ No providers trained in SU care |

| | | |
|---|---|---|
| **16.4** | **Do other clinics/departments at this health facility provide HIV-related services/care?** | ☐ Yes<br>☐ No **{SKIP TO Q16.6}** |

**16.5** **To what extent do each of the following clinics/departments at this health facility provide all aspects of HIV care and what is the extent of training in HIV care received by staff in that clinic/department?**

| | Clinic/department | HIV care provided in the clinic? | Providers trained in HIV care? |
|---|---|---|---|
| a | Tuberculosis (TB) clinic | ☐ All aspects of HIV care<br>☐ Some aspects of HIV care<br>☐ HIV care not provided **{SKIP TO Q16.4b}**<br>☐ Don't know **{SKIP TO Q16.4b}**<br>☐ Not applicable (no TB clinic) **{SKIP TO Q16.4b}** | ☐ All providers trained in HIV care<br>☐ Some providers trained in HIV care<br>☐ No providers trained in HIV care<br>☐ Don't know |
| b | Chronic/non-communicable disease (NCD) clinic (Diabetes, Hypertension, etc) | ☐ All aspects of HIV care<br>☐ Some aspects of HIV care<br>☐ HIV care not provided **{SKIP TO Q16.4c}**<br>☐ Don't know **{SKIP TO Q16.4c}**<br>☐ Not applicable (no NCD clinic) **{SKIP TO Q16.4c}** | ☐ All providers trained in HIV care<br>☐ Some providers trained in HIV care<br>☐ No providers trained in HIV care<br>☐ Don't know |
| c | General outpatient department (OPD) clinic | ☐ All aspects of HIV care provided in OPD clinic<br>☐ Some aspects of HIV care provided in OPD clinic<br>☐ HIV care not provided in OPD clinic **{SKIP TO Q16.6}**<br>☐ Don't know **{SKIP TO Q16.6}**<br>☐ Not applicable (no OPD clinic) **{SKIP TO Q16.6}** | ☐ All providers trained in HIV care<br>☐ Some providers trained in HIV care<br>☐ No providers trained in HIV care<br>☐ Don't know |

| | | |
|---|---|---|
| **16.6** | **At this health facility, has any new clinic or department been set up in the past 12 months to provide care for both HIV and other chronic conditions?** | ☐ Yes<br>☐ No **{SKIP TO Q16.8}** |

**16.7** **To what extent are the following services provided in the new HIV/chronic care clinic and what is the extent of training received by staff in the clinic/department?**

| | Service | Services provided in new clinic? | Providers in new clinic trained? |
|---|---|---|---|
| a | HIV care | ☐ All aspects of HIV care<br>☐ Some aspects of HIV care | ☐ All providers trained in HIV care<br>☐ Some providers trained in HIV care<br>☐ No providers trained in HIV care<br>☐ Don't know |
| b | Tuberculosis (TB) care | ☐ All aspects of TB care<br>☐ Some aspects of TB care<br>☐ TB care not provided in new clinic **{SKIP TO Q16.7c}**<br>☐ Don't know **{SKIP TO Q16.7c}** | ☐ All providers trained in TB care<br>☐ Some providers trained in TB care<br>☐ No providers trained in TB care<br>☐ Don't know |
| c | Hypertension (HTN) care | ☐ All aspects of HTN care<br>☐ Some aspects of HTN care<br>☐ HTN care not provided **{SKIP TO Q16.7d}**<br>☐ Don't know **{SKIP TO Q16.7d}** | ☐ All providers trained in HTN care<br>☐ Some providers trained in HTN care<br>☐ No providers trained in HTN care<br>☐ Don't know |
| d | Diabetes Mellitus (DM) care | ☐ All aspects of DM care<br>☐ Some aspects of DM care<br>☐ DM care not provided **{SKIP TO Q16.7e}**<br>☐ Don't know **{SKIP TO Q16.7e}** | ☐ All providers trained in DM care<br>☐ Some providers trained in DM care<br>☐ No providers trained in DM care<br>☐ Don't know |

**16.8** **For each of the systems and processes listed below, please indicate whether HIV-related care and non-HIV care use or share the same system or use different/separate systems.**

| | | |
|---|---|---|
| a | Is the same electronic medical record (EMR) or data system used for HIV care and for non-HIV care? | ☐ Yes (same system)<br>☐ No (different systems) |
| b | Is the same pharmacy/dispensing system used for both HIV and non-HIV medicines? | ☐ Yes (same system)<br>☐ No (different systems) |
| c | Is the same laboratory services system used for both HIV and non-HIV care? | ☐ Yes (same system)<br>☐ No (different systems) |
| d | Is the same logistics and supplies system used for both HIV and non-HIV care? | ☐ Yes (same system)<br>☐ No (different systems) |
| e | At this health facility, are HIV care and non-HIV care supervised by the same individual? | ☐ Yes (same supervisor)<br>☐ No (different supervisors) |

| 17 | WEATHER HAZARDS AND DISASTER PREPAREDNESS | |
|---|---|---|
| 17.1 | **To your knowledge, which of the following weather hazards have ever occurred in the area where this clinic is located?**<br><br>*Select all that apply or "None of the above".* | ☐ Heatwaves<br>☐ Droughts<br>☐ Extreme cold snaps (sudden, unexpectedly cold weather)<br>☐ Severe storms (cyclones/hurricanes, severe rainfall, blizzards, derechos, tornados, etc.)<br>☐ Flooding<br>☐ Wildfire or wildfire smoke<br>☐ Other<br>☐ None of the above **{SKIP TO Q17.5}** |
| 17.2 | **Has this clinic experienced any of the following weather hazards in the past 12 months?**<br><br>*Select all that apply or "None of the above".* | ☐ Heatwaves<br>☐ Droughts<br>☐ Extreme cold snaps (sudden, unexpectedly cold weather)<br>☐ Severe storms (cyclones/hurricanes, severe rainfall, blizzards, derechos, tornados, etc.)<br>☐ Flooding<br>☐ Wildfire or wildfire smoke<br>☐ Other<br>☐ None of the above **{SKIP TO Q17.5}** |
| 17.3 | **In the past 12 months, did this clinic experience any of the following disruptions because of hazardous weather?**<br><br>*Select all that apply or "None of the above".* | ☐ Structural damage to the clinic<br>☐ Loss of power/electricity for >2 days<br>☐ Loss of internet/telecommunication services for >2 days<br>☐ Disrupted/contaminated water supplies for >2 days<br>☐ Damage to transportation infrastructure (e.g. roadways)<br>☐ Damage to existing medical equipment, medication stocks or consumable supplies<br>☐ Disrupted supply chains (e.g. delays in receiving medical commodities/supplies)<br>☐ None of the above |
| 17.4 | **In the past 12 months, did your clinic experience any of the following challenges in providing HIV care because of hazardous weather?**<br><br>*Select all that apply or "None of the above"* | ☐ Staffing shortages or absences<br>☐ Disruptions in provision of care/medication to patients<br>☐ Disruptions in ability to contact/communicate with patients<br>☐ Disrupted data entry (e.g., health records, laboratory results)<br>☐ Reduced clinic hours<br>☐ Short-term clinic closure (<5 days)<br>☐ Longer-term clinic closure (≥5 days)<br>☐ Increased caseloads or patient transfers from other clinics affected by hazardous weather<br>☐ None of the above |
| 17.5 | **How does this clinic receive early warnings or other notifications about hazardous weather?**<br><br>*Select all that apply or "None of the above"* | ☐ Text/SMS alerts<br>☐ Telephone calls<br>☐ Email alerts<br>☐ Radio or television broadcasts<br>☐ Other announcements (e.g. bulletins or roadside signs)<br>☐ Don't know **{SKIP TO Q18.1}**<br>☐ This clinic does not receive early warnings of hazardous weather **{SKIP TO Q18.1}** |
| 17.6 | **Which of the following organizations provide warnings about hazardous weather conditions?**<br><br>*Select all that apply or "Don't know"* | ☐ Local government<br>☐ National authority<br>☐ Meteorological services (weather services)<br>☐ Non-governmental organizations<br>☐ Other<br>☐ Don't know |

| 18 | ACKNOWLEDGEMENTS |
|---|---|
| 18.0 | **Approximately how long (in hours) did it take you to complete this survey?** *Round to the nearest half-hour (for example, enter 1.5 for 90 minutes)* |
| 18.1 | **We would like to acknowledge clinic team members who participated in the completion of this survey.** If your team members would like their names included, please enter their full names (first name and SURNAME), separated by commas, so we can acknowledge their contribution. |

AUG-2026

**Thank you for your participation.**
